# The predictive value of interictal scalp EEG findings in aiding the detection of malformations of cortical development in temporal lobe epilepsy and impact on surgical planning

**DOI:** 10.1101/2021.05.08.21256883

**Authors:** Jeffrey W. Fuchs, Nathan A. Shlobin, Benjamin S. Hopkins, Zehra Husain, Michael B. Cloney, Evgeniya Tyrtova, Pue Farooque, Jessica W. Templer, S. Kathleen Bandt

## Abstract

**Background:** Complete resection of focal malformations of cortical development (MCD) has been recognized as crucial for the success of epilepsy surgery. However, many of these lesions escape detection using even state-of-the-art epilepsy protocol MRI imaging. This study evaluates the concurrence of radiographic and histopathologic findings of MCD in patients with refractory temporal lobe epilepsy (TLE) and defines the predictive value of EEG findings in the detection of MCD.

**Materials and Methods:** Pre-operative MRI, scalp VEEG, and post-operative surgical pathology reports from 34 consecutive patients treated for refractory TLE by surgical resection over a ten year period were included in analysis. Radiographic findings of MCD were correlated with histopathologic findings of MCD and compared against pre-operative interictal scalp EEG findings.

**Results:** 66.7% of focal cortical dysplasias (FCD) identified on pathology and all cases of mild MCD (mMCD) were missed on pre-operative MRI. The description of a rhythmic or continuous interictal abnormality on pre-operative VEEG corresponds to a sensitivity of 73.1% and a specificity of 62.5% in detecting either FCD or mMCD. Of the patients who had a radiographically occult FCD, 80% had either a continuous or rhythmic interictal abnormality described in the interpretation of their pre-operative VEEG.

**Conclusion:** This study highlights the high prevalence of MCDs in refractory TLE and the high rate of missed MCDs on pre-operative MRI. Findings here suggest that pre-operative scalp EEG may be able to provide additional information in the pre-operative detection of MCDs and therefore inform surgical decision making.

## Introduction

Epilepsy is one of the most common neurologic disorders which affects 1% of the population of the United States.^1^ Approximately 30% of patients with epilepsy do not achieve freedom from seizures with medication.^2^ Surgery may be offered to select patients in the treatment of medically refractory epilepsy. However, surgical cure rates still remain, at best, 70% despite the myriad diagnostic procedures including advanced neuroimaging with volumetric analysis.^3-6^

One common form of medically refractory focal-onset epilepsy is temporal lobe epilepsy (TLE). The pathologic findings in TLE have been well-studied and include hippocampal sclerosis, malformations of cortical development (MCD), tumors, and cavernous malformations.^7-12^ Barkovich *et. al*., described a broad definition of MCDs and introduced the notion of these representing a spectrum of developmental anomalies.^13^ Complete resection of dysplastic, focal MCDs, including “mild malformations of cortical development” (mMCD) and focal cortical dysplasia (FCD), has been recognized as crucial for the success of epilepsy surgery.^14-16^

With the advent of minimally invasive surgical approaches for medically refractory epilepsy including radiosurgery and laser thermal ablation, we now have the ability to target specific epileptogenic foci with great precision. Therefore, it has become even more critical to accurately identify the underlying etiology and location of the pathology causing a patient’s refractory epilepsy. In patients harboring an underlying pathology, such as an MCD, recognizing that pathology and considering it in the surgical treatment plan is critical to prevent the need for subsequent surgical procedures or a lifetime of continued epilepsy.

Although advances in diagnostic technology, including magnetic resonance imaging (MRI), have improved our ability to visualize lesions that are potential causes of refractory epilepsy, some lesions continue to escape our detection using standard visual interpretation techniques. The rate of radiographically occult MCDs has been reported to range between 25% and 45%.^14, 17-19^ Since the introduction of 3T MRI as an improvement upon the previously standard 1.5T MRI magnets, there has been only a marginal improvement in the detection of some MCDs.^20, 21^ However, the rate of occult MCDs overall remains stable, suggesting that visual inspection of MRI data alone should not be exclusively relied upon to identify these lesions.

The use of electroencephalography (EEG) in the diagnosis of TLE is well-established.^11^ However, the use of scalp EEG findings as a diagnostic predictor of cortical dysplasia is less certain.^22^ Cortical dysplasias, a subset of MCDs according to Barkovich’s classification scheme, have been shown to produce rhythmic epileptiform discharges on EEG that correspond to continuous epileptiform discharges on electrocorticography.^23, 24^ More recently, it has been hypothesized that non-epileptiform interictal abnormalities including focal irregular slowing seen on surface EEG is present in patients with subtle FCD.^25^ These findings suggest that the presence of rhythmic or continuous interictal abnormalities on surface EEG may be predictive of cortical dysplasia. Taken together, these descriptive patterns identified on scalp EEG may serve as supplementary data to suggest an underlying focal, dysplastic MCD may be to blame for a patient’s refractory epilepsy, regardless of MRI findings. This data can then inform subsequent surgical decision-making including the choice to use either intraoperative electrocorticography or the use of intracranial EEG monitoring. Both of these options can inform subsequent surgical planning to include a more extended surgical approach (temporal lobe resection) rather than a more focal surgical approach (laser ablation) if more broadly distributed abnormal findings are identified.

The purpose of this study is twofold. First, to evaluate the concurrence between radiographic findings of MCD and histopathological findings in patients who underwent surgery for refractory TLE. Second, to evaluate the predictive value of abnormalities on surface EEG in the identification of an underlying MCD.

## Methods

### Patient Population

We retrospectively identified all adult patients (>18 years of age) who were treated for medically refractory TLE by surgical resection in the Department of Neurosurgery of Northwestern University between January 2007 and December 2017. Surgical procedures performed include temporal lobectomy with or without amygdalohippocampectomy or resection of a specific seizure onset node (topectomy) within the temporal lobe (see Table 1 for breakdown by surgical procedure). Four patients were excluded who had a previous intracranial surgery for epilepsy, as well as those who did not have pre-operative video electroencephalography (VEEG) or epilepsy protocol MRI.

**Table 1:** Overview of surgical procedure, MRI findings and histopathological diagnosis from all patients undergoing temporal lobe surgery for refractory TLE between 2007 and 2017.

| Patient | Surgery | Epilepsy-Related MRI Findings | Pathology |
| --- | --- | --- | --- |
| 1 | Right TL and AH | Right Frontal Encephalomalacia | mMCD and HS |
| 2 | Right TL and AH | Encephalomalacia of Temporal Lobe. Left frontal and right parietal venous anomalies. | Temporal encephalomalacia with microgliosis |
| 3 | Left TL and AH | Left Mesial Temporal Sclerosis | mMCD and HS |
| 4 | Right TL and AH. Resection of Cavernous Angioma | Right Mesial Temporal Sclerosis. Right cerebral venous anomaly. | Cavernous angioma, mMCD, and HS |
| 5 | Resection of Epileptogenic Focus on Right Temporal Lobe | Right Temporal Lesion Consistent with Low-Grade Glioma. Hyperintense Lesion in the Right Anterior Temporal Cortex. | WHO grade II diffuse-type astrocytoma, protoplasmic variant |
| 6 | Right TL and AH | Right Mesial Temporal Sclerosis | Chaslin's gliosis and HS |
| 7 | Right TL and AH | Right Mesial Temporal Sclerosis | HS |
| 8 | Right TL and AH | None | FCD Type 2a |
| 9 | Right TL and AH | Right Temporal Cortical Lobe Dysplasia | FCD Type 2b and WHO Grade I ganglioglioma |
| 10 | Left TL and AH | Left Mesial Temporal Sclerosis | HS |
| 11 | Right TL and AH | None | FCD Type 2a and mMCD |
| 12 | Right TL and AH | Right Mesial Temporal Sclerosis | mMCD |
| 13 | Partial Right TL and Partial AH | None | mMCD |
| 14 | Left TL and AH | Left Mesial Temporal Sclerosis. Left Choroidal Fissure Cyst. | mMCD and HS |
| 15 | Left TL and AH | Left Mesial Temporal Sclerosis. | mMCD and HS |
| 16 | Right TL and AH | Right Mesial Temporal Sclerosis. Arachnoid cyst in the left middle cranial fossa. Sinus inflammatory changes in the posterior ethmoid air cells on the left with possible mucus retention. | mMCD and HS |
| 17 | Right TL and AH | Small vessel ischemic changes in the supratentorial subcortical and periventricular white matter. | mMCD and hippocampus with patchy gliosis |
| 18 | Right TL and Resection of Amygdala | Hyperintense cystic lesion extending through the floor of the anterior right middle cranial fossa and into the sphenoid bone. Thin rim of T2 isointense signal surrounding the cystic lesion which may represent dysplastic brain parenchyma. | Cerebral cortex and subcortical white matter with patchy gliosis. Focal mild astrogliosis |
| 19 | Right TL and AH | Right Mesial Temporal Sclerosis | mMCD and HS |
| 20 | Left TL and AH | Left Mesial Temporal Sclerosis | mMCD and HS |
| 21 | Left TL and AH | Volume loss involving the right hippocampus. | FCD Type 2a |
| 22 | Right TL and AH | Abnormal high FLAIR signal in the mesial right temporal lobe. Differential including cortical dysplasia, low grade glioma, post ictal edema, paraneoplastic syndrome, and encephalitis. | FCD Type 2a |
| 23 | Left TL and AH | None | Microinfarcts involving gray and white matter |
| 24 | Right TL and AH | None | FCD Type 2a and HS |
| 25 | Right TL and AH | None | FCD Type 2a and HS |
| 26 | Right TL and AH | Two nodular foci of heterotropic gray matter along the lateral aspect of the right temporal horn periventricular white matter. | FCD Type 2a |
| 27 | Right TL and AH | Blurring of gray-white differentiation within the anterior/inferior right temporal lobe. Finding could relate to cortical dysplasia. | FCD Type 2a and focal neuronal dropout consistent with ischemic injury in temporal lobe and hippocampus |
| 28 | Right TL and AH | Hyperintense T2/FLAIR signal within the right frontal white matter, left frontal white matter, and left parietal white matter. | FCD Type 2a |
| 29 | Resection of Epileptogenic Focus on Left Temporal Lobe | Cystic lesion identified within the left middle temporal gyrus with ongoing abnormal T2/FLAIR hyperintense signal surrounding it and extending into the left peritrial white matter where another smaller cystic process persists. | WHO Grade I Ganglioglioma arising from cortical and subcortical dysplasia (FCD) |
| 30 | Right TL and AH | Right amygdala found to be fuller than the left, possibly focal cortical dysplasia or low grade glioma. | FCD Type 2a and HS |
| 31 | Right TL and AH | None | FCD Type 2a and reactive gliosis of amygdala and hippocampus |
| 32 | Right TL and AH | Right mesial temporal sclerosis. Likely epidermal inclusion cysts. | HS |
| 33 | Left TL and AH | Left mesial temporal sclerosis | FCD Type 3a and HS |
| 34 | Right TL and AH | Right mesial temporal sclerosis | FCD Type 2a |

TL=temporal lobectomy, AH=amygdalohippocampectomy, FCD=focal cortical dysplasia, mMCD=mild malformation of cortical development, HS=hippocampal sclerosis.

Patients were identified to have temporal lobe epilepsy using diagnostic procedures including surface VEEG with or without the use of sphenoidal electrodes. All patients with diagnosed temporal lobe epilepsy had a pre-operative brain MRI performed in accordance with the epilepsy protocol at Northwestern University. While this protocol has evolved over time, all epilepsy protocol MRIs included axial and coronal T1, T2 and fluid attenuated inversion recovery (FLAIR) sequences at minimum. Tissue samples were obtained at the time of surgery and analyzed by expert neuro-pathologists at Northwestern University. Data collection for this study was approved by the Institutional Review Board (IRB) of Northwestern University (IRB#: STU00205860), and patient records were reviewed in accordance with the guidelines of the IRB. Patient consent was neither required nor sought because only deidentified data were collected.

### Study Design

Patient charts including pre-operative VEEG reports, pre-operative epilepsy protocol MRI reports, surgical reports, and surgical pathology reports were reviewed. Classification of EEG abnormalities was performed using reports from pre-operative VEEGs. Recognizing that the interpretation of EEG data can be subjective based on the experience and background of an individual reviewer, and that the language used in the description of EEG findings is based heavily on the reviewer’s training and institutional preferences, interictal abnormalities were classified according to common terms which were used to describe identified abnormalities. The descriptive terms which were identified for inclusion in this study were “continuous slowing,” “continuous sharp waves,” and “rhythmic slowing” as determined by board certified clinical epilepsy neurologists interpreting the VEEG study. A rhythmic interictal abnormality was identified by use of the term “rhythmic” in the body and/or impression of the study report and a continuous abnormality was identified by the term “continuous,” “frequent,” “abundant,” or having a frequency greater than one occurrence every thirty seconds as described in the body and/or impression of the study report. Example EEG images of interictal abnormalities which are related to the descriptor terms described above from the patients in the cohort can be seen in Figure 1. We specifically chose to use the initial impression of the treating epilepsy neurologist as documented in the clinical interpretation of the VEEG study rather than retrospectively reviewing the VEEG in the context of the pathologic findings in order to bolster the clinical utility of this finding on initial review of EEG data.

**Figure 1.**
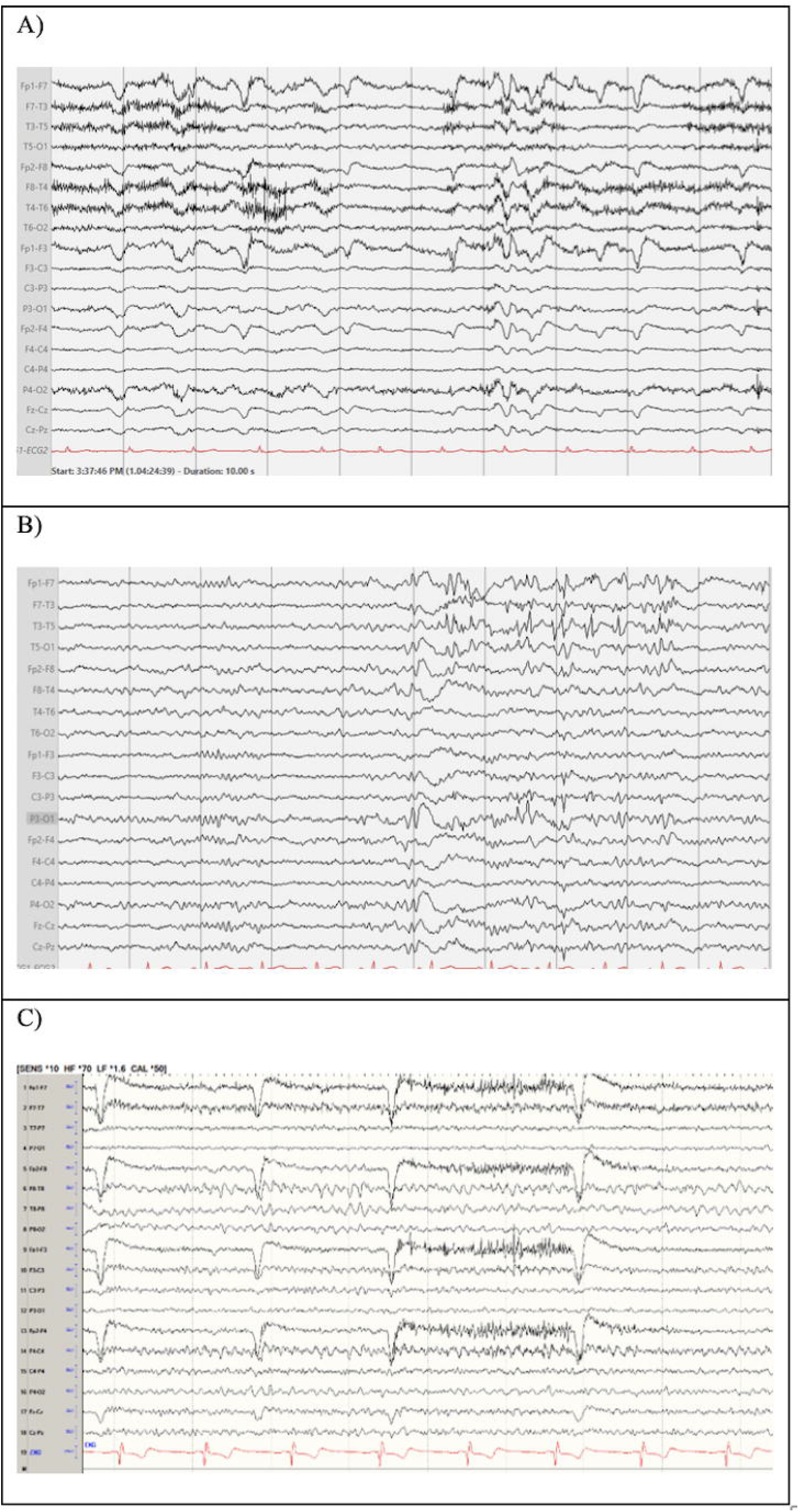
This figure shows example EEGs corresponding to the use of descriptive terms of A) continuous slowing, B) continuous sharp waves and, C) rhythmic slowing.

Findings from the pre-operative epilepsy protocol MRI reports were classified based on the interpretation of the reading neuroradiologist at the time of image acquisition. The classification scheme included identification of possible focal cortical dysplasia (FCD), hippocampal sclerosis (HS), infarct/lesion, or tumor. Again, the initial impression of the neuroradiologist as documented in the MRI report was selected for inclusion in our analysis.

Finally, findings from the surgical pathology report were classified based on the type of pathology identified. The classification scheme used was presence of FCD, mMCD, HS, temporal gliosis, infarct/encephalomalacia, or tumor. mMCDs were defined by normal cortex with excess ectopic or heterotopic neurons in the molecular layer or subcortical white matter according to the 2004 Palmini or 2011 Blümcke classification systems if not stated directly by the neuropathologist.^16, 26^

Classification of MRI, EEG, and pathology findings as described above was performed independently by two members of the study group (S.K.B and J.W.F). No discrepancies in classification of findings were identified.

Data are reported as counts for categorical variables and mean ± standard deviation for continuous variables. Microsoft Excel 2013 (Microsoft, Redmond, Washington) was used to conduct all statistical analyses.

## Results

### Demographics

We identified 34 patients who met inclusion criteria, 21 (60%) of whom were female, with an average age at time of surgery of 40.6 ± 13.2 years. The average age of onset of epilepsy was 20.9 ± 16.1 years and the average duration of seizures was 19.7 ± 16.8 years. A comprehensive review of the surgical procedures performed can be found in Table 1. Six patients had isolated FCD, two patients had isolated mMCD, one patient had both FCD and mMCD, eight patients had FCD co-occurring with another pathology, nine patients had mMCD co-occurring with another pathology, and eight patients had no evidence of MCD on histologic examination.

### MRI, Pathology, and VEEG Findings

Table 1 gives the MRI findings for each patient that were identified to be a possible cause of epilepsy along with the surgical pathology findings for each patient. The classification of VEEG reports, according to the interictal findings, compared against each category of histopathological finding can be found in Table 2.

**Table 2:** Rates of interictal EEG findings for a given histopathological diagnosis.

| Interictal EEG finding | Isolated<br>FCD<br>(n=10) | Isolated<br>mMCD<br>(n=3) | FCD +<br>mMCD<br>(n=1) | Isolated<br>HS (n=4) | HS + FCD<br>(n=4) | HS + mMCD<br>(n=8) | Other<br>(n=4) |
| --- | --- | --- | --- | --- | --- | --- | --- |
| Continuous Slowing (n=9) | 2 | 1 | 0 | 0 | 2 | 3 | 1 |
| Continuous Sharp Waves<br>(n=5) | 2 | 0 | 0 | 0 | 2 | 1 | 0 |
| Rhythmic Slowing (n=3) | 0 | 0 | 0 | 0 | 0 | 2 | 1 |
| Rhythmic Sharp Waves (n=0) | 0 | 0 | 0 | 0 | 0 | 0 | 0 |
| Continuous Slowing +<br>Continuous Sharp Waves<br>(n=1) | 1 | 0 | 0 | 0 | 0 | 0 | 0 |
| Continuous Slowing +<br>Rhythmic Slowing (n=2) | 2 | 0 | 0 | 0 | 0 | 0 | 0 |
| Continuous Slowing +<br>Rhythmic Sharp Waves (n=0) | 0 | 0 | 0 | 0 | 0 | 0 | 0 |
| Continuous Sharp Waves +<br>Rhythmic Slowing (n=2) | 1 | 0 | 0 | 1 | 0 | 0 | 0 |
| Continuous Sharp Waves +<br>Rhythmic Sharp Waves (n=0) | 0 | 0 | 0 | 0 | 0 | 0 | 0 |
| Rhythmic Slowing +<br>Rhythmic Sharp Waves (n=0) | 0 | 0 | 0 | 0 | 0 | 0 | 0 |
| None (n=12) | 2 | 2 | 1 | 3 | 0 | 2 | 2 |

FCD=focal cortical dysplasia, mMCD=mild malformation of cortical development, HS=hippocampal sclerosis. Other includes infarcts, encephalomalacia, or tumor.

### Identification of Malformations of Cortical Development on MRI versus Pathology

Of the five patients identified to have possible MCD on pre-operative MRI, all five had a confirmed diagnosis of FCD on histopathology. However, 10 patients (66.7%) were found to have FCD on pathology that was not identified on pre-operative MRI. None of the 12 confirmed cases of mMCD on pathology were identified on pre-operative MRI.

### EEG Findings as a Predictor of Malformations of Cortical Development

The finding of an interictal abnormality, including rhythmic or continuous slowing or sharp waves was seen in 22 of the 34 patients (64.7%) undergoing temporal lobe surgery for refractory epilepsy. A rhythmic or continuous interictal abnormality was seen in 12 (80%) of the 15 patients with FCD and 7 (58.3%) of the 12 patients with mMCD. Overall, a rhythmic interictal abnormality was seen in 19 (73%) of the 26 patients with MCD (FCD or mMCD). A rhythmic or continuous interictal abnormality was seen in 3 (37.5%) of the 8 patients without evidence of MCD.

The finding of a rhythmic or continuous interictal abnormality corresponds to a sensitivity of 80% and specificity of 47.4% for the presence of FCD and a sensitivity of 58.3% and a specificity of 31.8% for the presence of mMCD. The finding of a rhythmic or continuous interictal abnormality on VEEG to predict the presence of any type of MCD (FCD or mMCD) has a sensitivity of 73.1% and a specificity of 62.5%. The positive predictive value for the presence of MCD on pathology given the finding of a rhythmic or continuous interictal abnormality in this cohort was 86.4%.

A continuous interictal abnormality was more common on EEG than was the finding of a rhythmic interictal abnormality with 19 patients having continuous interictal slowing or sharp waves while only 7 patients had rhythmic interictal slowing. The finding of a continuous interictal abnormality alone corresponds to a sensitivity of 65.4% and specificity of 75% for the presence of any type of MCD (FCD or mMCD). The positive predictive value of any MCD in the presence of any continuous interictal abnormality in this cohort was 89.5%.

### EEG Findings as Predictor of FCD not Identified on MRI

Ten cases of histopathologically confirmed FCD were not identified using preoperative MRI (66.7%). Of these 10 radiographically-occult FCD, 8 patients had a finding of rhythmic or continuous interictal abnormality on EEG: four patients had continuous slowing, three patients had continuous sharp waves, and one had both continuous sharp waves and rhythmic slowing.

## Discussion

### Concordance Between MRI and Pathology

In this study, we sought to evaluate the concordance between radiographic findings of cortical dysplasia and histopathological findings in patients who underwent surgery for refractory TLE. This study found that 66.7% of FCDs found on pathology were missed on MRI. This rate of missed FCD is slightly higher than has been reported in previous studies.^14, 17-19^ This only supports the need to consider the possibility of MCD in all patients living with refractory epilepsy in order to minimize the chances of incomplete diagnosis. This report, together with others, further endorses the subtle nature of MCD on imaging and an inability to rely exclusively on standard radiologic analysis for their detection.

We also find that all 12 cases of histopathologically confirmed mMCD were missed on pre-operative MRI. This is not surprising given the high rate of missed overt FCD. However, according to the Barkovich classification of MCD, these should be included in the FCD spectrum, and therefore considered for their epileptogenic potential.^13^ Few studies have attempted to identify neuroimaging findings consistent with mMCD. In a single study investigating MRI findings in patients with mMCD, 29% of cases of mMCD were associated with lobar hypoplasia and atrophy on MRI and 29% of cases demonstrated blurring of the gray/white matter junction.^17^ Unfortunately, both of these features can suggest MCD in general and do not specifically suggest mMCD in particular.

### Usefulness of EEG Findings as a Predictor of Malformations of Cortical Development

The second goal of this study was to identify whether the use of descriptive terms identifying rhythmic or continuous interictal abnormalities, as seen on surface EEG, are predictive of MCDs. The percentage of patients with FCD having a rhythmic or continuous interictal abnormality in this study was found to be 80%. This is much larger than the percentage of patients with FCD who were found to have unifocal epileptiform discharges on scalp EEG as reported by Kim *et. al*. (42.7%) or rhythmic epileptiform discharges as reported by Gambardella *et. al*. (44%).^22, 23^ This higher rate of interictal findings is accounted for by our inclusion of both epileptiform and non-epileptiform abnormalities, unlike the aforementioned studies which only include epileptiform abnormalities.

In this study, we demonstrate that the description of a rhythmic or continuous interictal abnormality corresponds to a sensitivity of 73.1% and a specificity of 62.5% in detecting either FCD or mMCD. These interictal findings were present in 19 (73%) of the 26 patients with MCD. In comparison, a rhythmic or continuous interictal abnormality was seen in only 3 (37.5%) of the 8 patients without evidence of MCD. These results suggest that, although a finding of rhythmic or continuous interictal slowing on EEG does not definitively determine the presence of MCD, if a patient demonstrates either of these interictal abnormalities on EEG, it should be considered that the patient has an underlying MCD. In particular, the presence of a continuous interictal EEG finding (either slowing or sharp waves) is particularly suggestive of an underlying FCD with a sensitivity of 80%.

### Usefulness of EEG Findings as a Predictor of Missed FCD

We find that 80% of patients with missed FCD on pre-operative epilepsy protocol MRI had continuous or rhythmic interictal abnormalities on EEG. This finding, together with an 80% sensitivity in detecting FCD in the setting of a continuous interictal abnormality, suggests that there may be some advantage in considering these interictal EEG findings in the prediction of an underlying FCD that may escape detection on pre-operative MRI. Anticipating the possibility of a radiographically occult MCD may help inform surgical decision-making when considering laser ablation of the mesial temporal structures versus placement of intracranial electrodes to further localize seizure onset in a patient with subtle hippocampal findings on pre-operative MRI and seizures localizing to one temporal lobe on VEEG. Outcome data would suggest that temporal lobectomy yields a slightly more robust seizure freedom rate than does mesial temporal laser ablation but that patients with clear evidence of isolated hippocampal sclerosis perhaps are well-served by laser ablation given its minimally invasive nature.^27-29^ This somewhat reduced seizure freedom rate following laser ablation may suggest that in the absence of hippocampal sclerosis (and perhaps the presence of continuous and/or rhythmic interictal EEG findings as described here), occult MCD may contribute to some of the decreased efficacy of such a precise surgical approach.

### Limitations and the Evolution of Pathology Classification Systems

Given that terminology and classification of FCDs on histopathology has evolved over the course of this study it can be difficult to compare findings for a given subtype of FCD. Before 2011, the Palmini classification was most commonly used in histopathological classification of MCDs.^16^ However, classification of MCDs has shifted over time to the Blümcke classification.^26^ This change in use of classification systems may have led to differences in histopathologic classification and may account for the more frequent classification of pathology as FCD type 2a in Patients 21-34 in Table 1 who underwent surgery after 2011 when this classification was implemented. Another potential difference in classification can be seen in the pathology findings for Patient 33 which was identified as FCD type 3a and hippocampal sclerosis (HS). Given changes in classification over time, it is possible that this pathologic finding would have been previously described as mMCD and HS or FCD type 2a and HS. The results of this study are not significantly altered by this change in terminology, however, because the results were analyzed according to overall classification of FCD and not by individual FCD type.

Another limitation of this study is that 26 of the 34 cases were found to have an MCD on pathology and of these cases, 12 had dual pathology of MCD and HS. Although an interictal abnormality was seen in only 1 of the 4 patients with isolated HS compared to 10 of the 12 patients with MCD co-occurring with HS--which may suggest that the presence of an MCD is the true underlying cause of interictal abnormalities--because the comparison groups of cases with isolated HS and other pathologies were fewer in number, this makes the specificity of the study findings in identifying MCD less certain. In order to validate the study conclusions, the findings should be applied to additional data sets with larger numbers of non-MCD pathology.

## Conclusion

This study highlights the substantial prevalence of MCDs in medically refractory TLE and the high rate of missed MCDs using pre-operative MRI. This study also demonstrates the description of a continuous and/or rhythmic interictal abnormality (including slow or sharp wave discharges) on scalp EEG as predictive of the presence of an MCD in the temporal lobe. In particular, description of a continuous interictal abnormality is particularly suggestive of underlying MCD. Identification of one or more of these interictal findings may suggest an intracranial EEG procedure may be warranted prior to employing an ultra-precise minimally invasive surgical treatment option such as laser ablation.

## Data Availability

Characteristics of patients are visible in Table 2.

